# Significance of SARS-CoV-2 Specific Antibody Testing during COVID-19 Vaccine Allocation

**DOI:** 10.1101/2021.01.28.21250721

**Authors:** Akane B. Fujimoto, Inci Yildirim, Pinar Keskinocak

## Abstract

**Objective:** To assess the value of using SARS-CoV-2 specific antibody testing to prioritize the vaccination of susceptible individuals as part of a COVID-19 vaccine distribution plan when vaccine supply is limited.

**Methods:** A compartmental model was used to simulate COVID-19 spread when considering diagnosis, isolation, and vaccination of a cohort of 1 million individuals. The scenarios modeled represented 4 pandemic severity scenarios and various times when the vaccine becomes available during the pandemic. Eligible individuals have a probability p of receiving antibody testing prior to vaccination (p = 0, 0.25, 0.5, 0.75, and 1). The value of serology testing was evaluated by comparing the infection attack rate, peak infections, peak day, and deaths.

**Results:** The use of antibody testing to prioritize the allocation of limited vaccines reduces infection attack rates and deaths. The size of the reduction depends on when the vaccine becomes available relative to the infection peak day. The largest reduction in cases and deaths occurs when the vaccine is deployed before and close to the infection peak day. The reduction in the number of cases and deaths diminishes as vaccine deployment is delayed and moves closer to the peak day.

**Conclusions:** Antibody testing as part of the vaccination plan is an effective method to maximize the benefit of a COVID-19 vaccine. Decision-makers need to consider relative timing between the infection peak day and when the vaccine becomes available.

## INTRODUCTION

The effective deployment of safe and effective vaccines is a key intervention to control the spread of the coronavirus disease 2019 (COVID-19) pandemic by establishing herd immunity via immunization in a shorter time and without additional deaths and burden on healthcare systems. As of December 2020, there are 61 vaccine candidates against SARS-CoV-2, the novel coronavirus causing COVID-19, in the clinical development phase using different platforms (i.e., genetic, viral vector, protein-based, inactivated virus), efficacy, doses required, and storage considerations [1]. In the U.S., the vaccines developed by manufacturers Pfizer-BioNTech and Moderna were given a first emergency use authorization (EUA) on December 11 and December 18, 2020, respectively, which allows their distribution in the country [2, 3].

The number of vaccines available to countries in the next months will be limited due to manufacturing and logistic constraints. Due to the limited amount of vaccine supply, as well as that their distribution will be staggered, vaccine allocation guidelines are extremely important to ensure vaccine equity and effective distribution worldwide especially in resource-limited settings.

Various guidelines have been developed by governing organizations ahead of the vaccine distribution. The National Academies of Science, Engineering, and Medicine has developed a framework to assist policymakers when planning for vaccine allocation. This framework includes a phased allocation of vaccines that prioritizes essential workers and high-risk individuals while ensuring equity in the distribution [4]. Similarly, states in the U.S. are developing plans for vaccine distribution which includes vaccine storage, distribution, administration, community communication, safety, and capacity considerations. While these plans address and consider various aspects of the vaccine supply chain and administration, they do not consider the potential benefit that serologic testing and assessing SARS-CoV-2 specific antibody response can have while allocating vaccines.

Serologic tests can help determine the individuals who were previously infected with SARS-CoV-2 [5-8]. Although the duration of immunity to and the rate of SARS-CoV-2 reinfection is still to be determined, in the majority of the COVID-19 cases serum antibodies developed against SARS-CoV-2 specific proteins raise within 2-3 weeks of infection and are detectable for at least three to six months after exposure [9-11]. Even if previous infection does not provide an individual full immunity, prioritizing the limited number of vaccines to those who have not acquired any natural immunity could be beneficial. Due to this, the identification of individuals who have been previously infected including those who have not developed the distinctive symptoms, so recovered without knowing they had have had SARS-CoV-2 infection, could be a promising policy for a more effective allocation of vaccines while supply is limited. In addition, it is estimated that only 1 in every 7.7 cases or 13% of the total cases have been detected and reported with the majority of the true cases remaining undiagnosed [12]. Thus, it is vital to deploy mass serology testing to identify individuals who have recovered from the symptomatic or asymptomatic SARS-CoV-2 infection.

In this paper, we used a compartmental model to quantify the benefit of using serology testing to prioritize the vaccination of individuals who are susceptible as they have never been infected with COVID-19 before. We evaluated 5 scenarios with respect to the degree to which serology is used during vaccination. To model serology test utilization we assigned a probability *p* (0, 0.25, 0.5, 0.75, and 1) that an individual receives serology testing before vaccination. We tested vaccine administration under different disease spread intensity rates and time of vaccine availability and compared the *infection attack rate* (IAR), peak infections, peak day, and deaths.

## METHODOLOGY

We used an extended version of the susceptible-infected-recovered (SIR) compartmental model to capture the epidemiology and the natural history of the SARS-CoV-2 infection, diagnosis and isolation of infected individuals, and serology testing and vaccine allocation, as seen in Figure 1. The model parameters are shown in Table 1. The parameters were obtained from literature and guidelines from the US Centers for Disease Control and Prevention [13]. The simulated cohort size was 1 million individuals. The simulation starts on January 1^st^, 2020 with 1 infected symptomatic individual. We ran 10 replications of the simulation. The simulation was run for 550 days (approximately 1.5 years). A detailed description of the model formulation is found in the supplementary material. The simulation and modeling were done using the statistical software R [14].

**Table 1:** Model parameters.

| Parameter | Description | Value | Sources |
| --- | --- | --- | --- |
| $\beta_s$ | The transmission rate due to contact between susceptible and symptomatic infected subjects.<br>The transmission rate for asymptomatic infected subjects is 75% of $\beta_s$ ($\beta_a = 0.75\beta_s$) | High: 0.225<br>Medium-high: 0.195<br>Medium: 0.178<br>Low: 0.168 | Estimated from [13] |
| $P_o$ | Proportion of infected individuals that develop symptoms | 0.6 | [13, 15] |
| $\gamma$ | Rate of recovery of an infected subject | 1/14 | [21, 22] |
| $\mu$ | Infection fatality rate among symptomatic infected individuals. | 0.001 | Estimated from [16, 17] |
| $D_x$ | Rate of diagnosis and isolation of infected individuals | 0.015 | Estimated from [12] |
| $Q$ | Rate of self-isolation of symptomatic infected individuals without a diagnosis. | 0.05 | Assumption |

**Figure 1:**
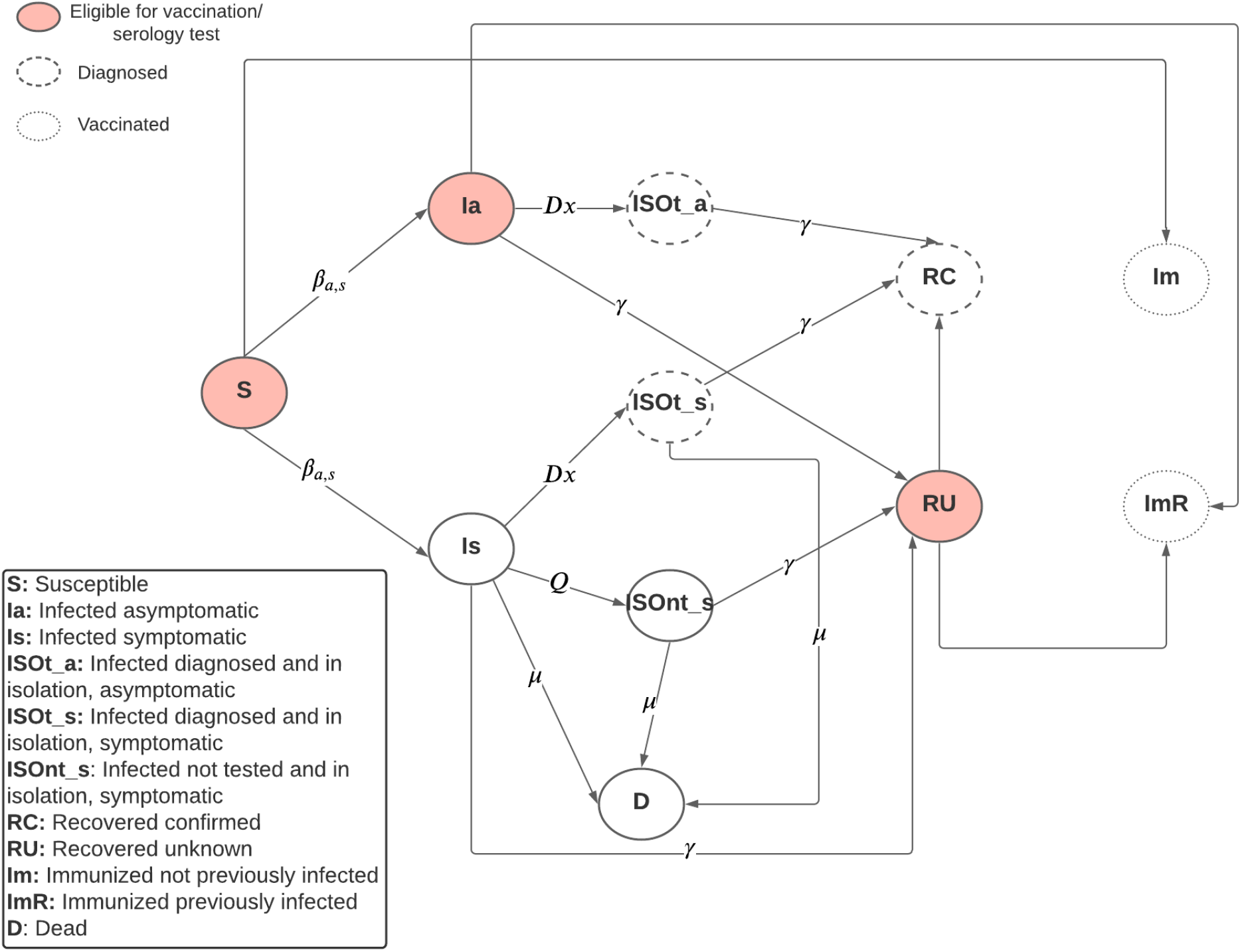
Compartmental model diagram.

### Infection

We extended the infected compartment of the basic SIR model to distinguish between symptomatic and asymptomatic infected individuals (Is and Ia) as they have different transmission rates. The infectiousness of asymptomatic infected individuals relative to symptomatic individuals is estimated to be 75% and 60% of infections are symptomatic [13, 15]. We considered 4 scenarios to illustrate a high, medium-high, medium, and low severity of the epidemic. The base reproductive number (R_0_) when interventions are not considered are 2.81, 2.43, 2.22, and 2.1 for the high, medium-high, medium, and low severity cases, respectively.

### Diagnosis, Isolation, Death, and Recovery

We added isolation compartments for the symptomatic and asymptomatic infected individuals who were diagnosed (ISOt_s and ISOt_a) and for the symptomatic infected individuals that decided to isolate voluntarily (ISOnt_s). In the isolation compartment, infected individuals have no contact with susceptible individuals, thus they do not transmit the disease. We assumed that infected individuals are diagnosed at a rate of 0.015 per day. This is equivalent to roughly 13% of the total cases to be diagnosed and confirmed by the end of the simulation, which follows practice [12]. Additionally, we assumed that symptomatic infected individuals would self-isolate at a rate of 0.05 per day. We assumed 100% compliance for isolation. The infection fatality rate among symptomatic individuals was 0.001, which was equivalent to a symptomatic infection fatality ratio of approximately 1.4% by the end of the simulation [16, 17]. We only considered mortality for infected symptomatic individuals. Infected individuals who recover move to the recovered compartment. We stratified this compartment to indicate if the infection was diagnosed (i.e., recovered and known infection, RC) or not (i.e., recovered and unknown infection, RU). We assumed that recovered individuals are immune, thus they can interact with infected individuals without risk of re-infection. When considering isolation, the effective reproductive number R_0_ becomes 1.76, 1.53, 1.39, and 1.32 for the high, medium-high, medium, and low epidemic severity cases, respectively.

### Serology Testing and Vaccine Distribution

We considered various times for when vaccines become available (within 7 months before and after the peak infection date during the pandemic in the baseline scenarios where vaccines are not available). We assumed that vaccines were available for 50% of the population (500,000 vaccine doses) and that the supply was uniformly distributed across 6 months which was equivalent to a daily supply of 2,748 units. Additionally, we reported results for when vaccines were available for 25% of the population. We assumed the serology tests employed were 100% sensitive and specific. In practice, sensitivity and specificity range from 66% to 97.8% and 96.6% to 99.7%, respectively, and depend on the technology used and the time between symptoms onset and when the blood sample is taken [18]. Vaccines are assumed to have an efficacy of 100%, therefore vaccinated individuals can interact with infected individuals without the risk of infection. The Pfizer-BioNTech and Moderna vaccines have a reported efficacy of 90% to 95% [19, 20]. Starting the day that the vaccines become available, we used the results of the compartmental model at the end of the day before to perform serology testing and vaccine administration. Eligible individuals for vaccination included the susceptible (S), infected asymptomatic (Ia), and recovered unknown (RU) populations. Individuals that are eligible for vaccination receive a serology test with probability *p* until vaccine capacity for the day is exhausted. We considered 5 values for *p* (0, 0.25, 0.5, 0.75, and 1) which indicates various degrees of serology testing usage from none to universal testing. If a serology test is given to the individual, they received a vaccine only if they are susceptible (S) or infected asymptomatic (Ia). Individuals in the recovered unknown (RU) compartment do not receive a vaccine and they are moved to the recovered known (RC) compartment. If a serology test is not given, individuals received a vaccine independently of their status. Individuals who received a vaccine are moved to one of the two compartments for immunized individuals depending on their previous status: Susceptible (S) individuals are moved to the immunized and not previously infected compartment (Im) while infected asymptomatic (Ia) and recovered unknown (RU) individuals are moved to the immunized but previously infected (ImR) compartment. After vaccines were allocated, the movement among the infected, isolated, dead, and recovered compartments occurred.

### Outcomes

The outcomes used to evaluate the scenarios and quantify the benefits of incorporating serology testing during vaccination plans include:

- Infection attack rate (IAR): Cumulative percentage of the population infected.
- Peak infections: Maximum number of new daily symptomatic and asymptomatic infections.
- Peak day: The day when the daily new symptomatic and asymptomatic infections are the highest.
- Deaths: Total number of deaths due to complications of COVID-19.

## RESULTS

The model outcomes of the baseline scenarios where no vaccines were available are displayed in Table 2. The results presented in this section correspond to the scenarios where the vaccine supply covers 50% of the population.

**Table 2:** Simulation output for the baseline scenarios.

| Transmission Rate | Base $R_0$ | Effective $R_0$ under isolation | Peak Day | IAR (%) | Peak Infections | Deaths |
| --- | --- | --- | --- | --- | --- | --- |
| High | 2.81 | 1.76 | June 4<sup>th</sup> 2020 | 72.33 | 13,596 | 5,992 |
| Medium-High | 2.43 | 1.53 | August 3<sup>rd</sup> 2020 | 60.43 | 7,951 | 5,006 |
| Medium | 2.22 | 1.39 | October 5<sup>th</sup> 2020 | 50.95 | 5,073 | 4,221 |
| Low | 2.10 | 1.32 | December 4<sup>th</sup> 2020 | 43.99 | 3,551 | 3,639 |

### Infection Attack Rate (IAR)

Figure 2 shows the IAR for the scenarios modeled. The highest IAR happens when the R_0_ is the highest and vaccination becomes available after the peak. The earlier the vaccine is deployed, the largest the reduction in the IAR. When vaccines are deployed too late (5 months or later after the peak), vaccine usage does not reduce the IAR compared to when vaccines are not available. The reduction in the IAR as a consequence of using serology tests to prioritize vaccination of susceptible individuals is the largest when the vaccines are deployed before and close to the infection peak day, as seen in Table 3. The largest reduction happens when the vaccine becomes available one or two months before the peak, depending on the scenario. In the scenario where the effective R_0_ is 1.53 (medium-high), the third-largest reduction happens when the vaccine is available three months ahead of the peak, followed by the same month as the peak. When the effective R_0_s are 1.39 (medium) and 1.32 (low), the third-largest reduction occurs when the vaccine becomes available the same month as the peak, followed by three months prior. On the other hand, when vaccines are deployed too early or too late (5 months or more months before or after the peak) using serology testing does not significantly affect the IAR. The reduction in the IAR due to serology testing depends on the scenario. For example, in the scenario where the R_0_ is 1.32 (low) and the vaccines become available two months before the peak, in October, using serology testing with *p*=1 results in a reduction of 9,138 cases while when *p*=0.5 results in a reduction of 4,451 cases compared to no serology testing (*p*=0).

**Table 3:** Percentage reduction in cumulative infections when compared to no serology testing a person receives a serology test with probability 0.25, 0.5, 0.75, and 1 when the vaccine is available for 50% of the population.

| R<sub>0</sub><br>(Baseline<br>Peak Day) | Vaccination<br>Timing | Months<br>between<br>vaccine<br>avail. and<br>peak | p |  |  |  |  |  |  |  |
| --- | --- | --- | --- | --- | --- | --- | --- | --- | --- | --- |
|  |  |  | 0.25 |  | 0.5 |  | 0.75 |  | 1 |  |
|  |  |  | Δ<br>Infections | Δ % | Δ<br>Infections | Δ % | Δ<br>Infections | Δ % | Δ<br>Infections | Δ % |
| 1.76<br>(Jun 4 -<br>2020) | May-20 | -1 | 6,456 | 1.18 | 13,495 | 2.46 | 21,459 | 3.91 | 31,080 | 5.66 |
|  | Jun-20 | 0 | 5,243 | 0.79 | 11,371 | 1.72 | 18,827 | 2.84 | 28,732 | 4.34 |
|  | Jul-20 | 1 | 1,970 | 0.28 | 4,542 | 0.64 | 8,152 | 1.15 | 13,748 | 1.94 |
|  | Aug-20 | 2 | 427 | 0.06 | 992 | 0.14 | 1,866 | 0.26 | 3,392 | 0.47 |
|  | Sep-20 | 3 | 70 | 0.01 | 183 | 0.03 | 358 | 0.05 | 667 | 0.09 |
|  | Nov-20 | 5 | 4 | 0.00 | 8 | 0.00 | 15 | 0.00 | 26 | 0.00 |
|  | Jan-21 | 7 | 1 | 0.00 | 1 | 0.00 | 1 | 0.00 | 1 | 0.00 |
| 1.53<br>(Aug 3 -<br>2020) | May-20 | -3 | 1,975 | 1.28 | 3,931 | 2.55 | 5,645 | 3.66 | 7,755 | 5.03 |
|  | Jun-20 | -2 | 4,359 | 1.45 | 8,753 | 2.90 | 13,166 | 4.37 | 17,998 | 5.97 |
|  | Jul-20 | -1 | 5,543 | 1.28 | 11,410 | 2.64 | 17,529 | 4.05 | 24,586 | 5.68 |
|  | Aug-20 | 0 | 4,686 | 0.88 | 9,869 | 1.86 | 15,791 | 2.97 | 22,991 | 4.33 |
|  | Sep-20 | 1 | 2,293 | 0.39 | 5,051 | 0.87 | 8,415 | 1.45 | 12,933 | 2.23 |
|  | Oct-20 | 2 | 772 | 0.13 | 1,743 | 0.29 | 3,007 | 0.50 | 4,826 | 0.81 |
|  | Nov-20 | 3 | 210 | 0.03 | 499 | 0.08 | 868 | 0.14 | 1,423 | 0.24 |
|  | Jan-21 | 5 | 18 | 0.00 | 42 | 0.01 | 70 | 0.01 | 115 | 0.02 |
| 1.39<br>(Oct 5 -<br>2020) | May-20 | -5 | 26 | 0.16 | 47 | 0.29 | 116 | 0.72 | 101 | 0.62 |
|  | Jul-20 | -3 | 1,436 | 1.14 | 2,624 | 2.09 | 3,701 | 2.95 | 4,923 | 3.92 |
|  | Aug-20 | -2 | 3,077 | 1.32 | 6,111 | 2.62 | 9,011 | 3.86 | 12,099 | 5.18 |
|  | Sep-20 | -1 | 4,242 | 1.24 | 8,461 | 2.47 | 12,866 | 3.76 | 17,746 | 5.19 |
|  | Oct-20 | 0 | 3,872 | 0.91 | 7,947 | 1.88 | 12,475 | 2.94 | 17,611 | 4.16 |
|  | Nov-20 | 1 | 2,410 | 0.51 | 5,103 | 1.08 | 8,190 | 1.73 | 11,929 | 2.52 |
|  | Dec-20 | 2 | 1,111 | 0.22 | 2,395 | 0.48 | 3,926 | 0.79 | 5,875 | 1.18 |
|  | Jan-21 | 3 | 430 | 0.09 | 931 | 0.18 | 1,551 | 0.31 | 2,317 | 0.46 |
| 1.32<br>(Dec 4 -<br>2020) | May-20 | -7 | - | - | - | - | - | - | - | - |
|  | Jul-20 | -5 | 113 | 0.47 | 134 | 0.56 | 202 | 0.84 | 314 | 1.31 |
|  | Sep-20 | -3 | 1,004 | 0.82 | 2,071 | 1.69 | 3,148 | 2.58 | 4,348 | 3.56 |
|  | Oct-20 | -2 | 2,286 | 1.12 | 4,451 | 2.19 | 6,723 | 3.31 | 9,138 | 4.49 |
|  | Nov-20 | -1 | 3,137 | 1.09 | 6,272 | 2.18 | 9,538 | 3.31 | 13,059 | 4.53 |
|  | Dec-20 | 0 | 2,966 | 0.84 | 6,067 | 1.71 | 9,325 | 2.63 | 13,023 | 3.68 |
|  | Jan-21 | 1 | 2,109 | 0.53 | 4,322 | 1.08 | 6,800 | 1.71 | 9,639 | 2.42 |

**Figure 2:**
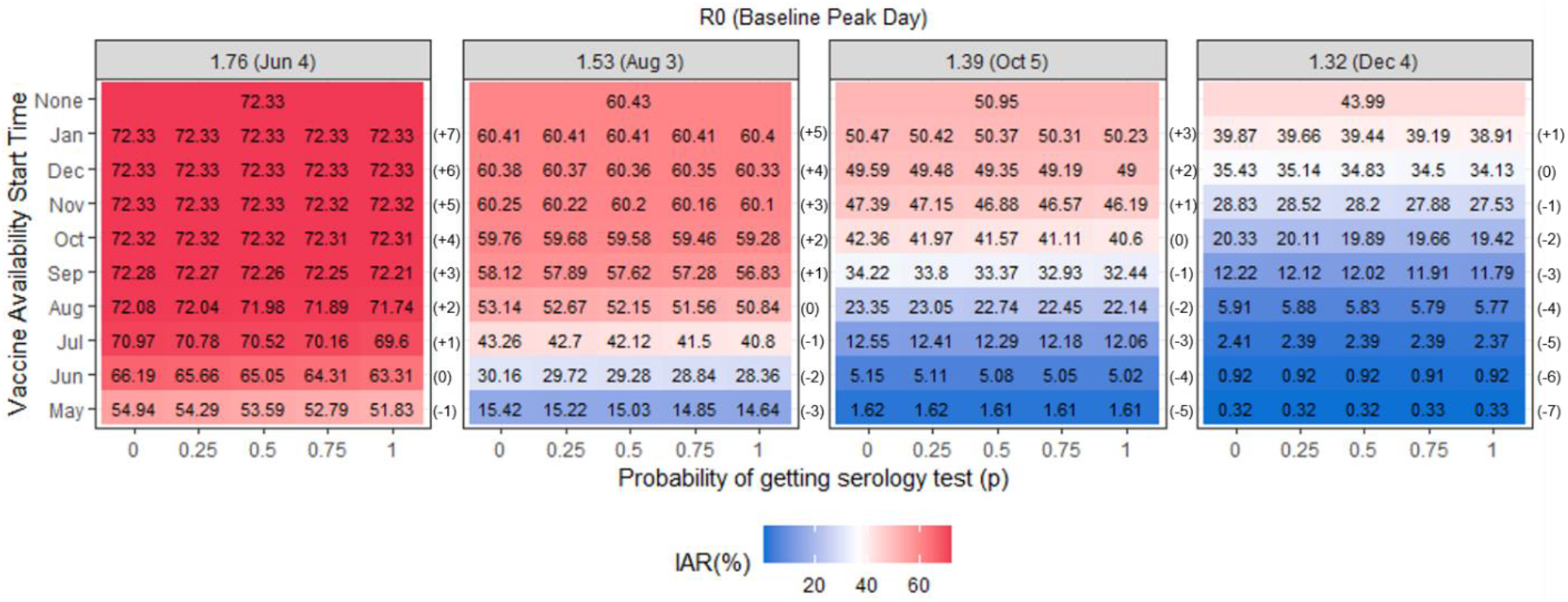
*Infection attack rate* for the scenarios evaluated when the vaccine is available for 50% of the population.

### Peak Infections and Peak Day

The timing of the vaccine availability has a significant effect on reducing the peak infections as seen in Figure 3. When vaccines are deployed before the baseline peak day it pushes the peak back (peak day is now earlier). When the vaccine is deployed after the baseline peak day, the peak infections and peak day remain unchanged. The use of serology testing helps to reduce the peak infections when the vaccine is deployed one or two months before the peak. The largest reduction of 177 new daily cases occurs when employing universal serology testing (*p* = 1), the R_0_ is 1.76 (high) and the vaccine becomes available one month before the peak. The peak day does not significantly change when serology testing is used and we evaluate scenarios with the same R_0_ and vaccine timing.

**Figure 3:**
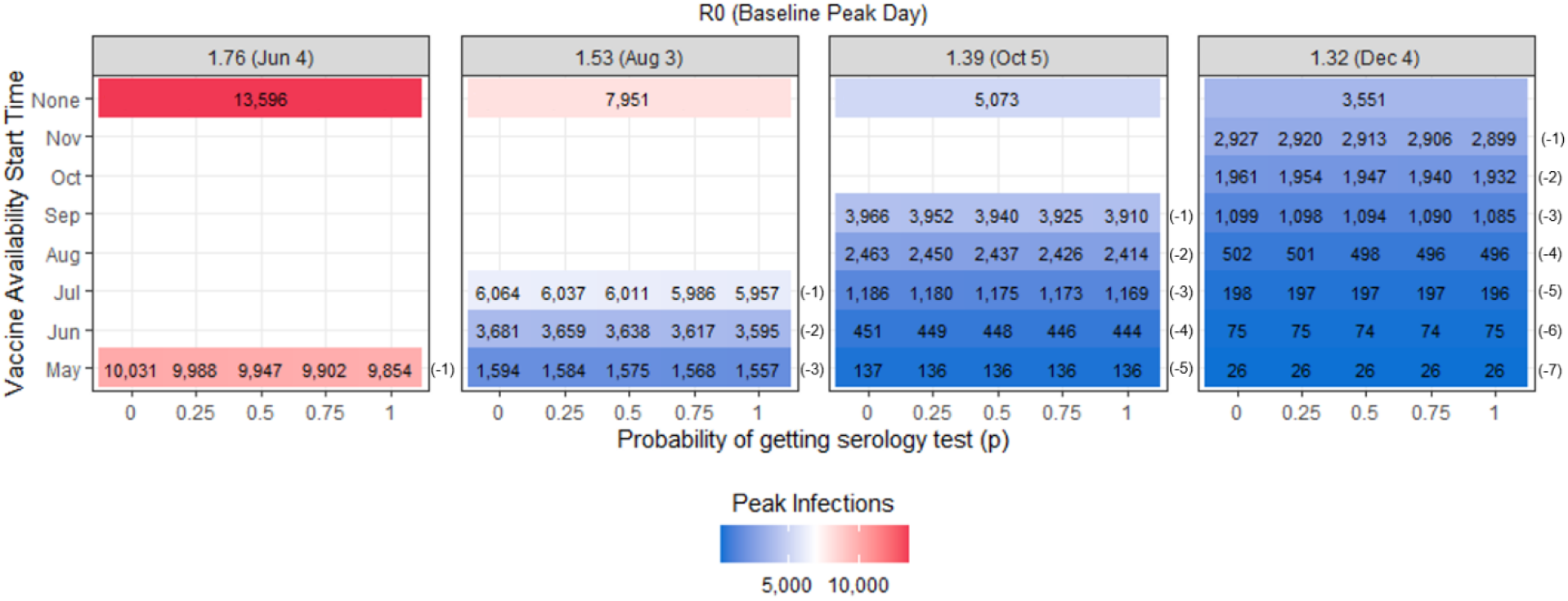
Peak infections for the scenarios evaluated when the vaccine is available for 50% of the population.

### Deaths

The earlier vaccines become available, the largest the reduction in deaths, as displayed in Figure 4. The use of serology testing has the largest reduction in deaths when the time of vaccine deployment is before and close to the baseline peak day, following the same pattern as to when evaluating IAR (Table 4). In the scenarios where the vaccine becomes available before and within three months of the peak, the reduction in deaths due to the use of serology ranges from 9 to 258 deaths. The largest reduction of 258 deaths occurs when employing universal serology testing (*p* = 1), the R_0_ is 1.76 (high) and the vaccine is available one month before the baseline peak.

**Table 4:** Percentage reduction in the number of deaths when compared to no serology testing a person receives a serology test with probability 0.25, 0.5, 0.75, and 1 when the vaccine is available for 50% of the population

| R<sub>0</sub><br>(Baseline<br>Peak Day) | Vaccination<br>Timing | Months<br>between<br>vaccine<br>avail. and<br>peak | p |  |  |  |  |  |  |  |
| --- | --- | --- | --- | --- | --- | --- | --- | --- | --- | --- |
|  |  |  | 0.25 |  | 0.5 |  | 0.75 |  | 1 |  |
|  |  |  | Δ Deaths | Δ % | Δ Deaths | Δ % | Δ Deaths | Δ % | Δ Deaths | Δ % |
| 1.76<br>(Jun 4 -<br>2020) | May-20 | -1 | 54 | 1.19 | 112 | 2.46 | 178 | 3.91 | 258 | 5.67 |
|  | Jun-20 | 0 | 43 | 0.78 | 94 | 1.71 | 156 | 2.85 | 238 | 4.34 |
|  | Jul-20 | 1 | 16 | 0.27 | 37 | 0.63 | 67 | 1.14 | 113 | 1.92 |
|  | Aug-20 | 2 | 3 | 0.05 | 8 | 0.13 | 15 | 0.25 | 28 | 0.47 |
|  | Sep-20 | 3 | 1 | 0.02 | 2 | 0.03 | 3 | 0.05 | 6 | 0.10 |
|  | Nov-20 | 5 | - | - | 1 | 0.02 | 1 | 0.02 | 1 | 0.02 |
|  | Jan-21 | 7 | - | - | - | - | - | - | - | - |
| 1.53<br>(Aug 3 -<br>2020) | May-20 | -3 | 16 | 1.25 | 32 | 2.51 | 46 | 3.60 | 64 | 5.01 |
|  | Jun-20 | -2 | 36 | 1.44 | 72 | 2.88 | 109 | 4.36 | 149 | 5.96 |
|  | Jul-20 | -1 | 45 | 1.26 | 94 | 2.62 | 145 | 4.05 | 203 | 5.66 |
|  | Aug-20 | 0 | 39 | 0.89 | 82 | 1.86 | 131 | 2.98 | 191 | 4.34 |
|  | Sep-20 | 1 | 19 | 0.39 | 42 | 0.87 | 70 | 1.45 | 107 | 2.22 |
|  | Oct-20 | 2 | 6 | 0.12 | 14 | 0.28 | 24 | 0.48 | 40 | 0.81 |
|  | Nov-20 | 3 | 2 | 0.04 | 4 | 0.08 | 7 | 0.14 | 12 | 0.24 |
|  | Jan-21 | 5 | - | - | 1 | 0.02 | 1 | 0.02 | 1 | 0.02 |
| 1.39<br>(Oct 5 -<br>2020) | May-20 | -5 | - | - | - | - | 1 | 0.75 | 1 | 0.75 |
|  | Jul-20 | -3 | 12 | 1.15 | 22 | 2.12 | 31 | 2.98 | 41 | 3.94 |
|  | Aug-20 | -2 | 26 | 1.34 | 51 | 2.64 | 75 | 3.88 | 101 | 5.22 |
|  | Sep-20 | -1 | 35 | 1.23 | 70 | 2.47 | 107 | 3.77 | 147 | 5.19 |
|  | Oct-20 | 0 | 32 | 0.91 | 66 | 1.88 | 103 | 2.94 | 146 | 4.16 |
|  | Nov-20 | 1 | 20 | 0.51 | 43 | 1.10 | 68 | 1.73 | 99 | 2.52 |
|  | Dec-20 | 2 | 9 | 0.22 | 20 | 0.49 | 33 | 0.80 | 49 | 1.19 |
|  | Jan-21 | 3 | 4 | 0.10 | 8 | 0.19 | 13 | 0.31 | 20 | 0.48 |
| 1.32<br>(Dec 4 -<br>2020) | May-20 | -7 | - | - | - | - | - | - | - | - |
|  | Jul-20 | -5 | 1 | 0.50 | 1 | 0.50 | 1 | 0.50 | 2 | 1.00 |
|  | Sep-20 | -3 | 9 | 0.89 | 18 | 1.78 | 26 | 2.57 | 36 | 3.56 |
|  | Oct-20 | -2 | 19 | 1.13 | 37 | 2.20 | 56 | 3.32 | 76 | 4.51 |
|  | Nov-20 | -1 | 26 | 1.09 | 52 | 2.18 | 79 | 3.31 | 108 | 4.52 |
|  | Dec-20 | 0 | 24 | 0.82 | 50 | 1.70 | 77 | 2.62 | 108 | 3.68 |
|  | Jan-21 | 1 | 18 | 0.55 | 36 | 1.09 | 57 | 1.73 | 80 | 2.42 |

**Figure 4:**
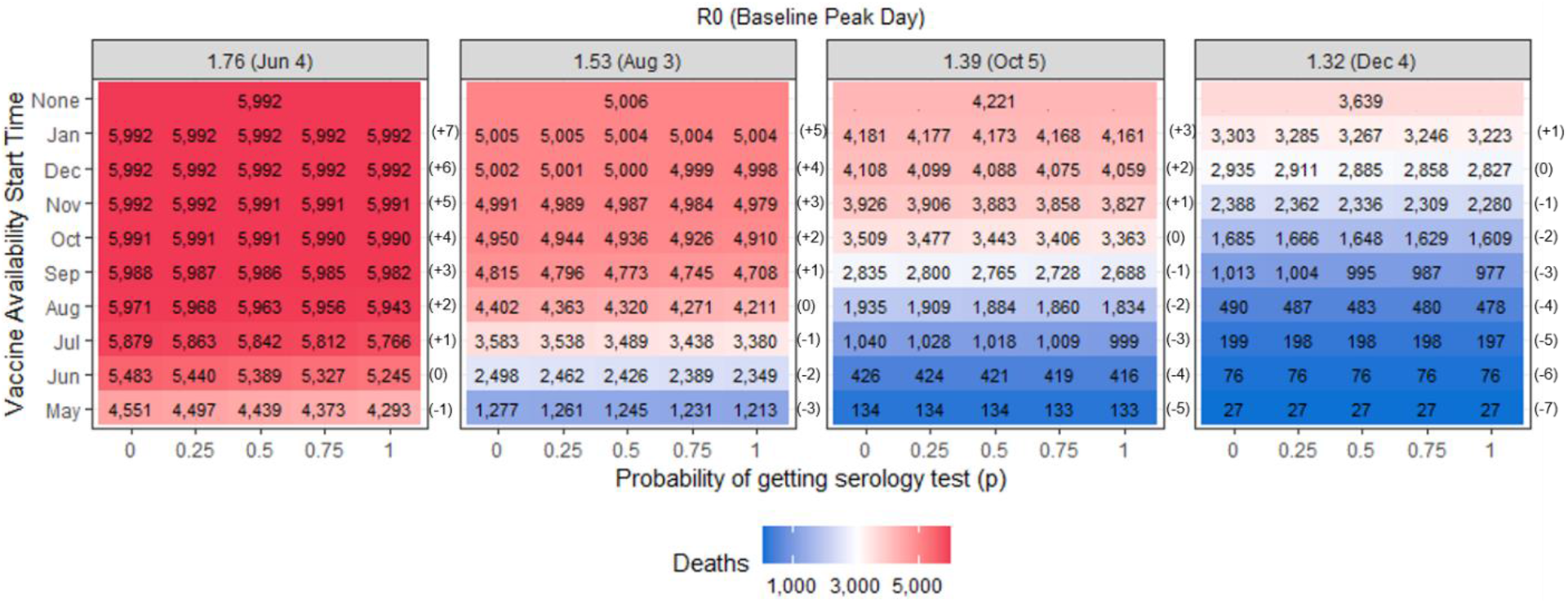
Total number of deaths for the scenarios evaluated when the vaccine is available for 50% of the population.

The results when the vaccine supply covers 25% of the population can be found in the supplementary material. When the supply decreases, we observed a similar pattern as to when it covered 50% of the population. The use of serology testing reduces IAR and deaths when the vaccines are deployed before and close to the infection peak. The largest reduction in IAR and deaths as a consequence of using serology testing occurs when the vaccine becomes available three months before the peak, followed by two and one months prior, in that order.

## DISCUSSION

The deployment of vaccines is an effective intervention to reduce the spread of a pandemic by limiting the number of susceptible individuals that can get infected by the disease safely and timely. Vaccine supply will be limited as the pandemic affects countries worldwide. Therefore, it is important to evaluate policies that can provide us with effective and fair allocation guidelines for vaccines. In this paper, we evaluated the use of serology testing to prioritize the allocation of the limited supply of vaccines to susceptible individuals, as opposed to individuals who have previously contracted the disease and are immune to some extent but are unaware of their status. The value of using serology testing comes from the information that it provides regarding the status of the individuals concerning the existence of a previous infection at the time of vaccination.

Serology testing programs have been already deployed in large-scale geographic surveys as well at the local level to estimate the true prevalence of COVID-19 [23-27]. Additionally, serological testing has been previously explored as a way to deploy a non-medical intervention called “shield immunity” where recovered individuals substitute susceptible individuals to avoid shutting down businesses and services while limiting the spread of the disease [28].

The simulation results show that vaccination has the largest reduction in IAR and deaths when it is deployed as early as possible. When the vaccine is deployed too late, after 5 months or later, vaccines do not significantly reduce the IAR and deaths. Employing serology testing to prioritize administering the vaccines to those who are fully susceptible reduces the IAR and the deaths. The largest reductions occur when the vaccine becomes available before and close to the peak day of the pandemic. The benefit gained by using serology testing diminishes as the vaccines are deployed too early or late. When comparing the impact of serology testing among scenarios with the same R_0_, the largest reduction in IAR and deaths occurs when the vaccines become available one or two months before the peak day, depending on the scenario, in the case where the vaccine supply covers 50% of the population (Tables 3 and 4). When the vaccines are available relatively early or late (more than 5 months before or after the peak), the reduction in the IAR and deaths when using serology testing is marginal. Ideally, vaccine deployment should occur as early as possible, and the use of serology testing should be evaluated depending on how early or late the vaccine becomes available relative to the disease progression.

We tested scenarios where serology testing was deployed at various intensities. The results of the model show that when universal serology testing is not viable (i.e., *p=*1), adopting an approach where only a fraction of the eligible individuals receive serology tests is still effective. When the supply of the vaccine decreases (i.e., vaccine covers 25% of the population instead of 50%), serology testing offers similar benefits regarding the reduction in IAR and deaths.

The earliest the vaccine is introduced, the largest the reduction will be observed in the peak infections. A reduction in peak infections implies a decrease in the number of daily new cases. This is important to consider since access to healthcare services can become impaired if the number of infected individuals seeking medical attention is greater than the available capacity. The use of serology testing reduces the peak infections when the vaccine is deployed one or two months before the peak day (Figure 3). In all other scenarios, serology testing does not significantly affect the infection peak. Similarly, serology testing does not significantly affect the resulting peak day.

The results of the modeling show that the use of serology testing to decide vaccine allocation priority groups reduces the IAR and deaths. Since the magnitude of the reduction depends on the number of serology tests employed and the relative time between the infection peak day and when vaccination becomes available, policymakers must consider the trade-off between the cost of the serology testing and the marginal benefit of its implementation given the state of the pandemic.

### Limitations

The results presented are limited by the restriction of our model and the assumptions made for the parameters explained in the methodology section. We use an extended SIR model which assumes homogenous mixing of the population. We do not stratify the population by age and therefore cannot account for age-dependent model parameters, such as the infection fatality ratio [13]. The infection transmission rates used in the model are static. In reality, these rates change over time depending on the interventions put in place and community compliance [29, 30]. We assumed that serology tests are 100% sensitive and specific and that vaccines are 100% effective, which do not reflect the real-world circumstances. We did not specify if the serology tests measure IgG or IgM or IgA antibodies. Additionally, we modeled vaccine distribution as a one-dose vaccine. The vaccines available in the market require two-doses separated by 21-28 days [31, 32]. In our model assuming a one-dose vaccine is equivalent to simulating a perfect administration of a two-dose vaccine (e.g., nobody misses the second dose). Although the limitations listed, the model proposed captures the pandemic dynamics and the allocation of vaccines using serology testing. The model provides us with a simple and accurate representation of the benefits of using serology as part of the vaccination process.

## CONCLUSIONS

The use of serology testing as part of the vaccine implementation process is an effective method to maximize the benefit of a COVID-19 vaccine. It provides a tool to identify and deliver the limited supply of the vaccine to a larger portion of the population that may be more susceptible to SARS-CoV-2 infection and as a result increases the likelihood of success of COVID-19 vaccination efforts. Identifying and prioritizing vaccination of those individuals who are susceptible to infection reduces the IAR and deaths. The magnitude of the reduction depends on the relative timing between the infection peak day and the time when vaccines become available. Policymakers need to evaluate the cost-effectiveness of deploying serology tests based on the current spread of the pandemic in their community and the timeline for the vaccine distribution.

## Data Availability

Only publicly available data has been used for this paper and it is cited in the manuscript.

## Acknowledgments

The authors are grateful to representatives from the Georgia Department of Public health for sharing information and expertise regarding vaccine distribution.

