## supplementary material for "Significance of SARS-CoV-2 Specific Antibody Testing during COVID-19 Vaccine Allocation"

### A. Compartmental Model

We extended the susceptible-infected-recovered (SIR) compartmental model to include additional compartments to capture the epidemiology and the natural history of the SARS-CoV-2 infection, diagnosis and isolation of infected individuals, and serology testing and vaccine allocation. We have 5 main compartments which are further divided into various sub-compartments.

1. Susceptible
   a. S: Susceptible

2. Infected
   a. Ia: Infected Asymptomatic
   b. Is: Infected Symptomatic
   c. ISOt_a: Infected diagnosed and in isolation, asymptomatic
   d. ISOt_s: Infected diagnosed and in isolation, symptomatic
   e. ISOnt_s: Infected not tested and in isolation, symptomatic

3. Recovered
   a. RC: Recovered confirmed
   b. RU: Recovered unknown

4. Immunized
   a. Im: Immunized not previously infected
   b. ImR: Immunized previously infected

5. Dead
   a. D: Dead

The parameters used in the model are:

- $\beta_s$, $\beta_a$: Transmission rate of due to contact between susceptible and symptomatic/asymptomatic infected subjects.
- γ: Rate of recovery of an infected subject.
- Po: Proportion of infected individuals that develop symptoms.
- μ: Infection fatality rate among symptomatic infected individuals.
- Dx: Rate of diagnosis and isolation of infected individuals
- Q: Rate of self-isolation of symptomatic infected individuals without a diagnosis.

The system of differential equations displayed below governs the movement of the population from/to the susceptible, infected, recovered, and dead compartments. The Euler method is used to solve the system of differential equations using the R package *deSolve*. The time unit used is days and the step size is 1 day.

- **Susceptible:**
  $\frac{dS}{dt} = -\beta_a(S)(I_a) - \beta_s(S)(I_s)$ (- new infections)
- **Infected Asymptomatic:**
  $\frac{dI_a}{dt} = (1 - P_o)[\beta_a(S)(I_a) + \beta_s(S)(I_s)] - \gamma I_a - dx(I_a)$ (+ new infections – recovered – diagnosed)
- **Infected Symptomatic:**
  $\frac{dI_s}{dt} = (P_o)[\beta_a(S)(I_a) + \beta_s(S)(I_s)] - \gamma I_s - dx(I_s) - Q(I_s) - \mu(I_s)$
  (+ new infections – recovered – diagnosed – undiagnosed quarantined - deaths)
- **Diagnosed Isolated Asymptomatic:**
  $\frac{dISO_{t\_a}}{dt} = dx(I_a) - \gamma ISO_{t\_a}$ (+ diagnosed – recovered)
- **Diagnosed Isolated Symptomatic:**
  $\frac{dISO_{t\_s}}{dt} = dx(I_s) - \gamma ISO_{t\_s} - \mu(ISO_{t\_s})$ (+ diagnosed – recovered – deaths)
- **Undiagnosed Isolated Symptomatic:**
  $\frac{dISO_{nt\_s}}{dt} = Q(I_s) - \gamma ISO_{nt\_s} - \mu(ISO_{nt\_s})$ (+ undiagnosed quarantined – recovered - deaths)
- **Recovered Confirmed:**
  $\frac{dRC}{dt} = \gamma(ISO_{t\_s} + ISO_{t\_a})$ (+ recovered)
- **Recovered Unknown:**
  $\frac{dRU}{dt} = \gamma(I_a + I_s + ISO_{nt\_s})$ (+ recovered)
- **Dead:**
  $\frac{dD}{dt} = \mu(I_s + ISO_{t\_s} + ISO_{nt\_s})$ (+ deaths)

When the vaccines become available at the specified times, individuals are moved to the immunized compartment according to the serology and vaccine allocation process described below.

**Serology and Vaccine Allocation Process**

1. We take the resulting values of the compartmental model from the day before as input.
2. We create a queue of individuals that are eligible for vaccination (i.e., individuals in the susceptible (S), infected asymptomatic (Ia), and recovered unknown (RU) compartments).
    - The order in the queue is random and people are selected with equal probability.

3. We toss a coin (0 < *toss* < 1) for the first individual in the queue.
4. We allocate a serology test or not if:
    - $toss \leq p$: Individual gets serology test.
    - $toss > p$: Individual doesn't get serology test.
5. We allocate a vaccine or not depending on if the individual received a serology test or not, per step 4:

    5a. If the individual gets a serology test and the individual's compartment is:

    - Susceptible (S): Vaccine is allocated, the individual is moved to the immunized not previously infected (Im) compartment.
    - Infected and asymptomatic (Ia): Vaccine is allocated, the individual is moved to the immunized previously infected (ImR) compartment.
    - Recovered unknown (RU): Vaccine is not allocated, the individual is moved to the recovered confirmed (RC) compartment.

    5b. If the individual does not get a serology test and the individual's compartment is

    - Susceptible (S): Vaccine is allocated, the individual is moved to the immunized not previously infected (Im) compartment
    - Infected and asymptomatic (Ia): Vaccine is allocated the individual is moved to the immunized previously infected (ImR) compartment.
    - Recovered unknown (RU): Vaccine is allocated, the individual is moved to the immunized previously infected (ImR) compartment.
6. Repeat steps 1-5 until daily vaccine capacity is allocated

**Reproductive Number ($R_0$) Estimation**

The base $R_0$ when no interventions are considered is:

$$R_{0,base} = P_0\left(\frac{\beta_s}{\gamma + \mu}\right) + (1 - P_0)\left(\frac{\beta_a}{\gamma}\right)$$

The effective $R_0$ when diagnosis and isolation are considered is:

$$R_{0,effective} = P_0\left(\frac{\beta_s}{\gamma + \mathrm{Dx} + \mathrm{Q} + \mu}\right) + (1 - P_0)\left(\frac{\beta_a}{\gamma + \mathrm{Dx}}\right)$$

### B. Results when vaccine supply covers 25% of the population

The results when the vaccine supply covers 25% of the population follow the same patterns as when the vaccine is available for 50% of the population.

#### Infection Attack Rate (IAR)

Figure B1 shows the IAR for the scenarios modeled. The reduction in the IAR as a consequence of using serology tests to prioritize vaccination of susceptible individuals is the largest when the vaccines are deployed before and close to the infection peak day, as seen in Table B1. The largest reduction happens when the vaccine becomes available three months prior before the peak, followed by two and one months prior, in that order.

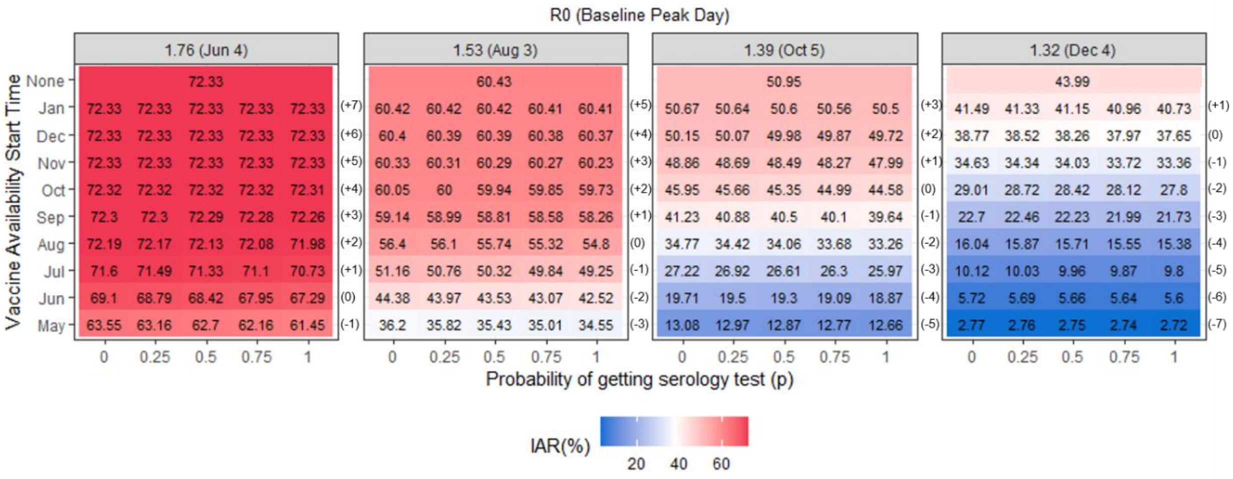


**Figure B1:** *Infection attack rate* for the scenarios evaluated when the vaccine is available for 25% of the population.

**Table B1:** Percentage reduction in cumulative infections when compared to no serology testing a person receives a serology test with probability 0.25, 0.5, 0.75, and 1 when the vaccine is available for 25% of the population.

| $R_0$ (Baseline Peak Day) | Vaccination Timing | Months between vaccine avail. and peak | p | | | | | | | |
|---|---|---|---|---|---|---|---|---|---|---|
| | | | 0.25 | | 0.5 | | 0.75 | | 1 | |
| | | | Δ Infections | Δ % | Δ Infections | Δ % | Δ Infections | Δ % | Δ Infections | Δ % |
| 1.76 (Jun 4 - 2020) | May-20 | -1 | 3,893 | 0.61 | 8,484 | 1.33 | 13,919 | 2.19 | 21,022 | 3.31 |
| | Jun-20 | 0 | 3,070 | 0.44 | 6,789 | 0.98 | 11,506 | 1.67 | 18,114 | 2.62 |
| | Jul-20 | 1 | 1,139 | 0.16 | 2,723 | 0.38 | 4,959 | 0.69 | 8,672 | 1.21 |
| | Aug-20 | 2 | 248 | 0.03 | 597 | 0.08 | 1,146 | 0.16 | 2,161 | 0.30 |
| | Sep-20 | 3 | 46 | 0.01 | 114 | 0.02 | 226 | 0.03 | 432 | 0.06 |
| | Nov-20 | 5 | 2 | 0.00 | 5 | 0.00 | 9 | 0.00 | 17 | 0.00 |
| | Jan-21 | 7 | - | - | - | - | - | - | 1 | 0.00 |
| 1.53 (Aug 3 - 2020) | May-20 | -3 | 3,780 | 1.04 | 7,696 | 2.13 | 11,831 | 3.27 | 16,426 | 4.54 |
| | Jun-20 | -2 | 4,097 | 0.92 | 8,491 | 1.91 | 13,178 | 2.97 | 18,612 | 4.19 |
| | Jul-20 | -1 | 4,024 | 0.79 | 8,401 | 1.64 | 13,247 | 2.59 | 19,067 | 3.73 |
| | Aug-20 | 0 | 3,057 | 0.54 | 6,638 | 1.18 | 10,788 | 1.91 | 16,006 | 2.84 |
| | Sep-20 | 1 | 1,468 | 0.25 | 3,297 | 0.56 | 5,574 | 0.94 | 8,767 | 1.48 |
| | Oct-20 | 2 | 497 | 0.08 | 1,137 | 0.19 | 2,001 | 0.33 | 3,258 | 0.54 |
| | Nov-20 | 3 | 138 | 0.02 | 330 | 0.05 | 581 | 0.10 | 967 | 0.16 |
| | Jan-21 | 5 | 8 | 0.00 | 22 | 0.00 | 43 | 0.01 | 76 | 0.01 |
| 1.39 (Oct 5 - 2020) | May-20 | -5 | 1,016 | 0.78 | 2,107 | 1.61 | 3,093 | 2.37 | 4,211 | 3.22 |
| | Jul-20 | -3 | 3,023 | 1.11 | 6,120 | 2.25 | 9,184 | 3.37 | 12,508 | 4.60 |
| | Aug-20 | -2 | 3,549 | 1.02 | 7,166 | 2.06 | 10,978 | 3.16 | 15,118 | 4.35 |
| | Sep-20 | -1 | 3,529 | 0.86 | 7,327 | 1.78 | 11,365 | 2.76 | 15,889 | 3.85 |
| | Oct-20 | 0 | 2,873 | 0.63 | 6,041 | 1.31 | 9,571 | 2.08 | 13,714 | 2.98 |
| | Nov-20 | 1 | 1,672 | 0.34 | 3,641 | 0.75 | 5,897 | 1.21 | 8,740 | 1.79 |
| | Dec-20 | 2 | 774 | 0.15 | 1,700 | 0.34 | 2,808 | 0.56 | 4,263 | 0.85 |
| | Jan-21 | 3 | 293 | 0.06 | 644 | 0.13 | 1,094 | 0.22 | 1,694 | 0.33 |
| 1.32 (Dec 4 - 2020) | May-20 | -7 | 50 | 0.18 | 205 | 0.74 | 308 | 1.11 | 488 | 1.76 |
| | Jul-20 | -5 | 869 | 0.86 | 1,650 | 1.63 | 2,490 | 2.46 | 3,266 | 3.23 |
| | Sep-20 | -3 | 2,433 | 1.07 | 4,764 | 2.10 | 7,150 | 3.15 | 9,725 | 4.28 |
| | Oct-20 | -2 | 2,889 | 1.00 | 5,805 | 2.00 | 8,810 | 3.04 | 12,024 | 4.15 |
| | Nov-20 | -1 | 2,961 | 0.85 | 5,995 | 1.73 | 9,169 | 2.65 | 12,696 | 3.67 |
| | Dec-20 | 0 | 2,456 | 0.63 | 5,067 | 1.31 | 7,903 | 2.04 | 11,113 | 2.87 |
| | Jan-21 | 1 | 1,566 | 0.38 | 3,336 | 0.80 | 5,309 | 1.28 | 7,608 | 1.83 |

**Peak Infections and Peak Day**

The resultant peak infections are displayed in Figure B2. The largest reduction in peak infections when using serology testing happens when $R_0$ is high (1.76) and equals 120 cases. The peak day does not significantly change when serology testing is used and we evaluate scenarios with the same $R_0$ and vaccine timing.

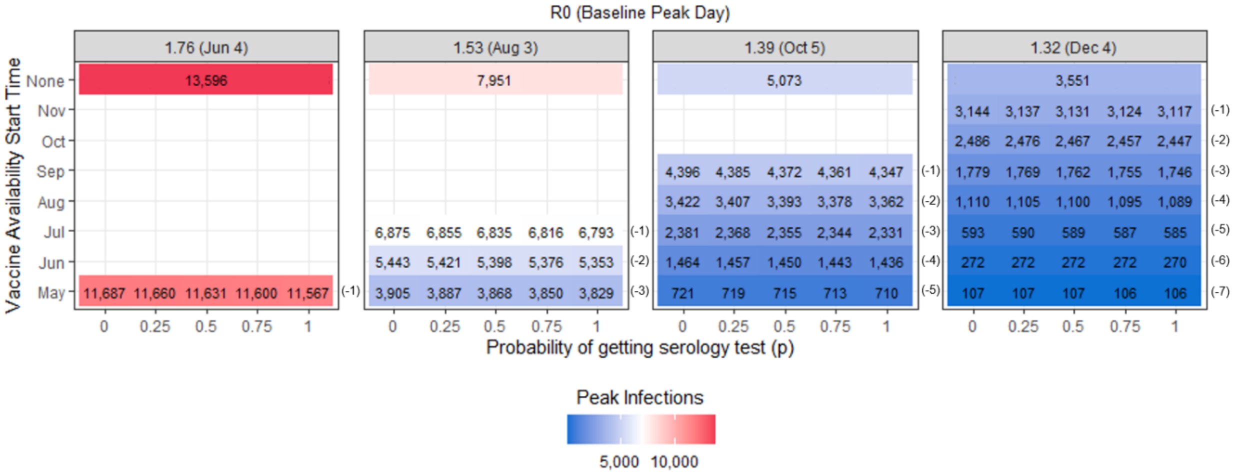


**Figure B2:** Peak infections for the scenarios evaluated when the vaccine is available for 25% of the population.

**Deaths**

Figure B3 shows the total number of deaths for the scenarios evaluated. The use of serology testing has the largest reduction in deaths when the time of vaccine deployment is before and close to the baseline peak day, following the same pattern as to when evaluating IAR (Table B2).

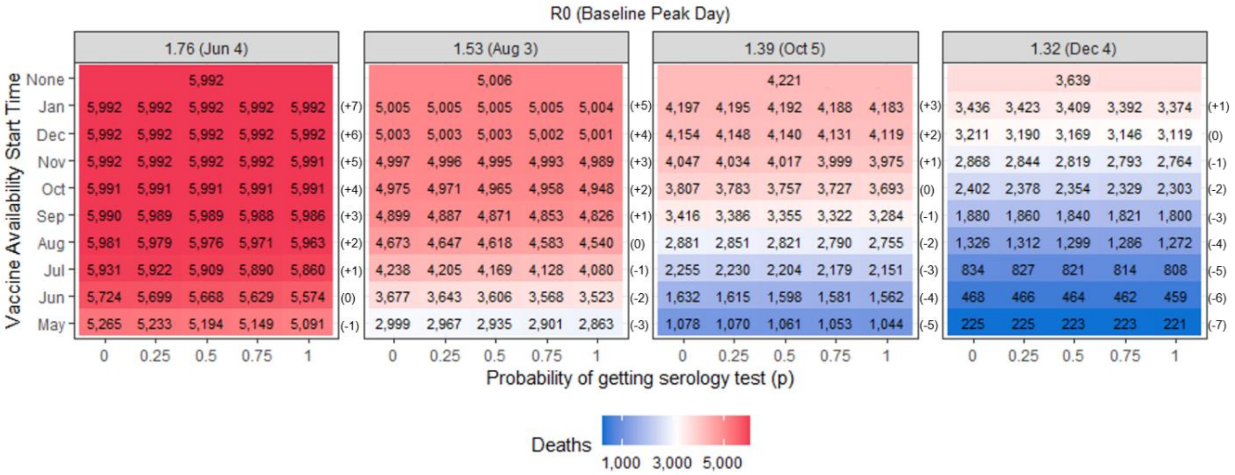


**Figure B3:** Total number of deaths for the scenarios evaluated when the vaccine is available for 25% of the population.

**Table B2:** Percentage reduction in the number of deaths when compared to no serology testing a person receives a serology test with probability 0.25, 0.5, 0.75, and 1 when the vaccine is available for 25% of the population.

| $R_0$ (Baseline Peak Day) | Vaccination Timing | Months between vaccine avail. and peak | p | | | | | | | |
|---|---|---|---|---|---|---|---|---|---|---|
| | | | 0.25 | | 0.5 | | 0.75 | | 1 | |
| | | | Δ Deaths | Δ % | Δ Deaths | Δ % | Δ Deaths | Δ % | Δ Deaths | Δ % |
| 1.76 (Jun 4 - 2020) | May-20 | -1 | 32 | 0.61 | 71 | 1.35 | 116 | 2.20 | 174 | 3.30 |
| | Jun-20 | 0 | 25 | 0.44 | 56 | 0.98 | 95 | 1.66 | 150 | 2.62 |
| | Jul-20 | 1 | 9 | 0.15 | 22 | 0.37 | 41 | 0.69 | 71 | 1.20 |
| | Aug-20 | 2 | 2 | 0.03 | 5 | 0.08 | 10 | 0.17 | 18 | 0.30 |
| | Sep-20 | 3 | 1 | 0.02 | 1 | 0.02 | 2 | 0.03 | 4 | 0.07 |
| | Nov-20 | 5 | - | - | - | - | - | - | 1 | 0.02 |
| | Jan-21 | 7 | - | - | - | - | - | - | - | - |
| 1.53 (Aug 3 - 2020) | May-20 | -3 | 32 | 1.07 | 64 | 2.13 | 98 | 3.27 | 136 | 4.53 |
| | Jun-20 | -2 | 34 | 0.92 | 71 | 1.93 | 109 | 2.96 | 154 | 4.19 |
| | Jul-20 | -1 | 33 | 0.78 | 69 | 1.63 | 110 | 2.60 | 158 | 3.73 |
| | Aug-20 | 0 | 26 | 0.56 | 55 | 1.18 | 90 | 1.93 | 133 | 2.85 |
| | Sep-20 | 1 | 12 | 0.24 | 28 | 0.57 | 46 | 0.94 | 73 | 1.49 |
| | Oct-20 | 2 | 4 | 0.08 | 10 | 0.20 | 17 | 0.34 | 27 | 0.54 |
| | Nov-20 | 3 | 1 | 0.02 | 2 | 0.04 | 4 | 0.08 | 8 | 0.16 |
| | Jan-21 | 5 | - | - | - | - | - | - | 1 | 0.02 |
| 1.39 (Oct 5 - 2020) | May-20 | -5 | 8 | 0.74 | 17 | 1.58 | 25 | 2.32 | 34 | 3.15 |
| | Jul-20 | -3 | 25 | 1.11 | 51 | 2.26 | 76 | 3.37 | 104 | 4.61 |
| | Aug-20 | -2 | 30 | 1.04 | 60 | 2.08 | 91 | 3.16 | 126 | 4.37 |
| | Sep-20 | -1 | 30 | 0.88 | 61 | 1.79 | 94 | 2.75 | 132 | 3.86 |
| | Oct-20 | 0 | 24 | 0.63 | 50 | 1.31 | 80 | 2.10 | 114 | 2.99 |
| | Nov-20 | 1 | 13 | 0.32 | 30 | 0.74 | 48 | 1.19 | 72 | 1.78 |
| | Dec-20 | 2 | 6 | 0.14 | 14 | 0.34 | 23 | 0.55 | 35 | 0.84 |
| | Jan-21 | 3 | 2 | 0.05 | 5 | 0.12 | 9 | 0.21 | 14 | 0.33 |
| 1.32 (Dec 4 - 2020) | May-20 | -7 | - | - | 2 | 0.89 | 2 | 0.89 | 4 | 1.78 |
| | Jul-20 | -5 | 7 | 0.84 | 13 | 1.56 | 20 | 2.40 | 26 | 3.12 |
| | Sep-20 | -3 | 20 | 1.06 | 40 | 2.13 | 59 | 3.14 | 80 | 4.26 |
| | Oct-20 | -2 | 24 | 1.00 | 48 | 2.00 | 73 | 3.04 | 99 | 4.12 |
| | Nov-20 | -1 | 24 | 0.84 | 49 | 1.71 | 75 | 2.62 | 104 | 3.63 |
| | Dec-20 | 0 | 21 | 0.65 | 42 | 1.31 | 65 | 2.02 | 92 | 2.87 |
| | Jan-21 | 1 | 13 | 0.38 | 27 | 0.79 | 44 | 1.28 | 62 | 1.80 |
